# Proposing a Weight-Based Expectation-Maximization Algorithm for Estimating Discrete-Time Markov Transition Probability Matrices with a Proof-of-Concept Example in Health Technology Assessment

**DOI:** 10.1101/2025.01.02.25319899

**Authors:** Marc Bollée, Abhirup Dutta Majumdar

## Abstract

Discrete-time Markov cohort-state transition models are now well-established as the preferred choice of analysts across application areas including health technology assessment. This preference arises out of its relative intuition and its capability to strike a fine balance between complex disease pathways, statistical precision, and parsimony although being criticized by a wide variety of stakeholders. Transition probability matrices (TPMs) are the “heart and soul” of such models responsible for estimating patient dispositions. However, estimating such TPMs comes with its own set of challenges. In some situations, the transition data may be censored such that the health state of a patient is unknown for multiple time steps before the next observation or data immaturity especially in rare diseases. Craig and Sendi proposed the expectation-maximization (EM) algorithm using uniform weights as a solution for unequal estimation intervals for partially observed data. However, this typically comes at the cost of increased within-state output variations with no optimization technique available in the literature.

The objective of this paper is to explore an optimized weighted version of the original EM algorithm, that aims to estimate the set of weights which minimizes the uncertainty of the estimated TPM against a target objective function. The weighting reduces the uncertainty of the estimate by considering the difference in temporal sparsity of the data when there are missing time steps. Further, we demonstrate the applicability of this weighting method using a fictitious cost-effectiveness model with our approach, showing a fine but definitive change over the original approach.

## 1. Introduction

The popularity of Markov cohort-state-transition models (MCSTMs) has not subsided over the decades across several application contexts, considering its inherent strength of balancing the clinical complexity of disease progression with statistical precision, and parsimony^1,2^. Transition probability matrices (TPM) are the most important determinant of the patient disposition across states of any MCSTM throughout the model horizon. Unfortunately, major data limitations often blockade such estimations. It is not uncommon for them to be performed using multiple sources of data input, unequal observation intervals, and missing transitions that contribute to the estimation problem. Srivastava et al 2021^3^ recently discussed the challenges modelers face when estimating TPs for cost-effectiveness modeling (CEM). In case of unequal observation intervals, Craig and Sendi^4^ have proposed an elegant approach that leverages full information at discrete time points which employs an uniformly weighted expectation-maximization (EM) algorithm. However, this typically may increase within state-transition variance and may impact the probabilistic sensitivity analysis (PSA)^5^ unnecessarily. Therefore, in this paper, we aim to propose an optimized (“weighted”) version of the EM algorithm and demonstrate its applicability using a fictitious toy example.

## 2. Methods

Let us assume that the input data is a set of transition count matrices observed over *s* time steps, denoted by *C*^(*s*)^ from one or multiple sources. During the E-step, using the current transition probability estimate 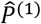, each *C*^(*s*)^ is decomposed into the estimated “most probable” set of 1-step transitions which resulted in *C*^(*s*)^. This 1-step estimate is denoted *Ĉ* ^(*s*→1)^. Then, during the M-step, 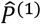 is recomputed based on these estimated 1-step count matrices. Now there are multiple sources of data to estimate 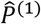, namely *C*^(1)^, *Ĉ* ^(2→1)^, ⋯, *Ĉ* ^(*S*→1)^. At this point, Craig and Sendi^4^ recommend adding these estimates together to create a single combined estimate which is shown in equation (1).

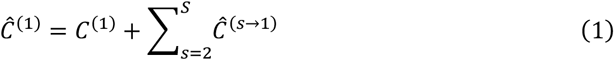

Subsequently, the maximum likelihood estimate of the transition probability matrix is obtained simply by the row-normalization of the combined 1-step count matrix. This updated estimate, 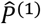, is then used again to repeat the E-step. This iterative process stops when an arbitrary level of convergence is achieved.

### 2.1 Rationale behind Weighting

The hypothesis is that although each of the different *Ĉ* ^(*s*→1)^ matrices are estimating the same 1-step counts, namely *C*^(1)^, they do not necessarily have identical distributions for the following two reasons. Firstly, the data generation and subsequent E-step process is different for each of the *Ĉ* ^(*s*→1)^. Secondly, each of the *C*^(*s*)^ matrices may be obtained from different data sources with varying observation intervals or can be prone to missingness. For example, one might pool multiple sources of clinical data to estimate the natural history of progression between disease states^6^. Therefore, taking an unweighted average may be suboptimal for minimizing the output variance. The proposed combination step is given by equation (2).

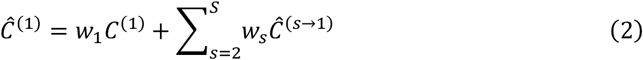

This is motivated by a simple result from the theory of best linear unbiased estimators^7^. To illustrate, estimating the mean of several normal random variables with identical mean but different variances, the best linear estimator of the mean is the one for which the weights are inversely proportional to the sample’s variance, as given in equation (3). To draw a further analogy, meta-analysis is founded on the idea of weighting effect sizes when pooling across studies to account for statistical heterogeneity^8^. However, the weighting effect from the perspective of estimating TPMs using the EM approach can only handle the inherent within state transition variability from similar data sources. For example, given a set of *n* independent observations 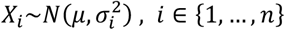, and 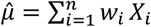, such that ***w*** ∈ Δ^3^ (*i. e*. 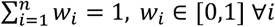, then one can prove:

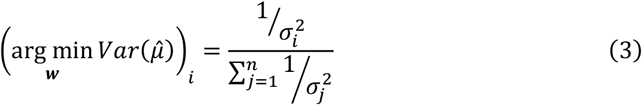

Note that the weighting does not affect 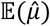 implicitly, since all *X*_i_ estimate the same parameter, namely *μ*. The calculation for normally distributed variables can be carried out using the method of Lagrange multipliers, however, the calculation for the weights of the estimated 1-step count matrices is not so simple. Firstly, there is no single function for the variance of a matrix. Secondly, whichever “variance” measure is used for the matrices (e.g. the sum of variances of the components) will have a complex relationship with the weights used. To alleviate this analytical complexity, a computational approach is taken. For a given function of the TPM, 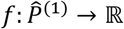, a grid search is performed to find the optimal weights that minimize the variance of the output of *f*. This function can be the sum of the variance of all components of 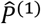, or the sum of the lengths of the 95% confidence intervals, or even the incremental cost-effectiveness ratio (ICER).

Craig and Sendi use Efron’s bootstrap^4,9,10^ to estimate the uncertainty around 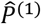. From the estimate, as many count matrices are generated at random as were in the original dataset. Then, the distribution of bootstrapped estimates 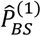, obtained by applying the EM algorithm to these artificial count matrices, allows us to approximately compute variance and 95% confidence interval of 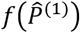. The process-flow is explained in *Figure 1*.

**Figure 1:**
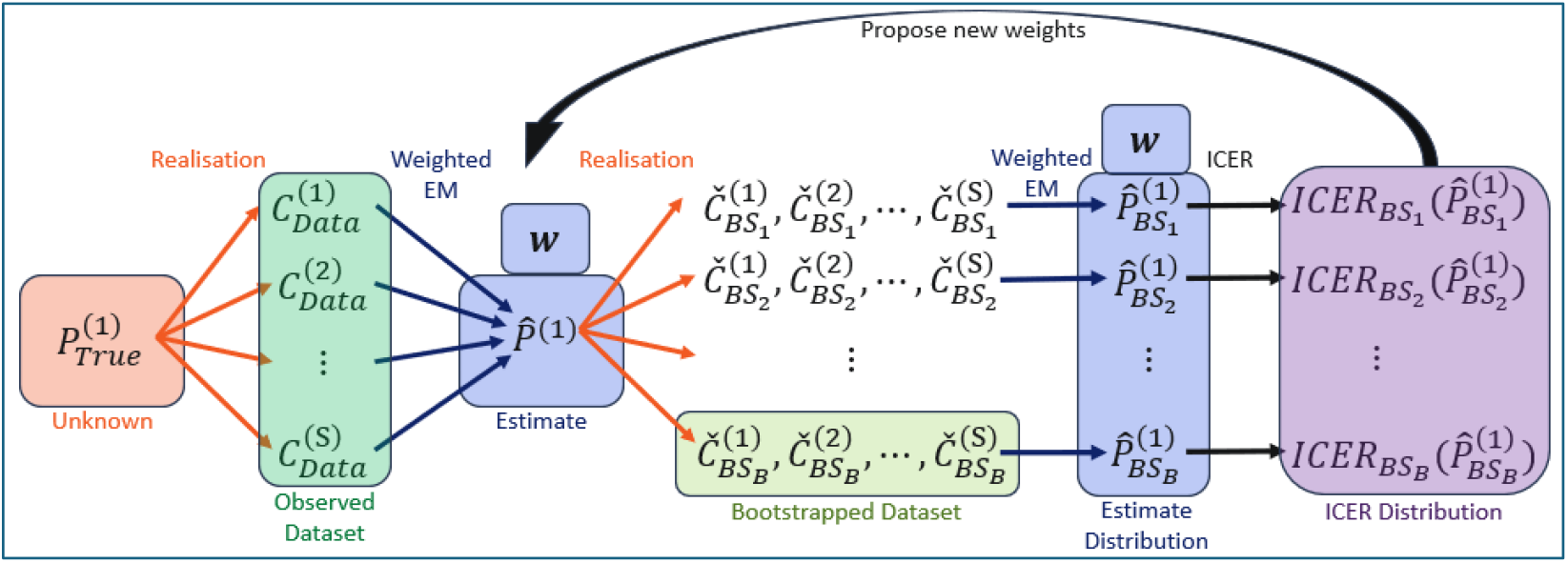
Process flow for bootstrapping of the transition kernel estimate

## 3. A Proof-of-Concept Example Using a 3-state Markov Cohort-based Transition Model

This example will illustrate how to adjust the weights to obtain a tighter confidence interval around the ICER of a fictitious new treatment “SuperDrug” which is prescribed as a primary prophylaxis as well as a subsequent treatment when diagnosed. The lifetime patient journey is modeled by a discrete-time Markov model with 3 hypothetical health states, “Healthy”, “Unhealthy”, and “Death” with a cycle length of 1 month (see Figure 2). The 1-month probability of transitioning between health states 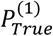 is unknown for SuperDrug, but out of a target sample population, 1-month transition counts have been observed, *C*^(1)^. However, on occasion, some patients failed to report their health state for one or two months in a row, resulting in 2-month and 3-month transition count matrices *C*^(2)^ and *C*^(3)^ respectively. The lack of reporting is assumed to be independent of the current health state (i.e. missing at random).

**Figure 2:**
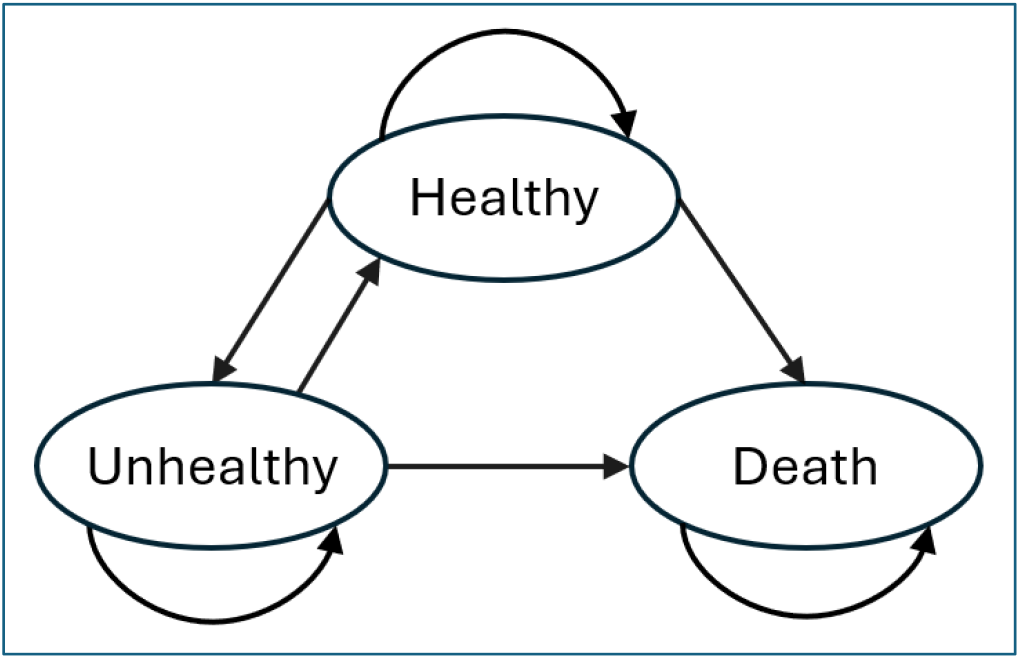
Disease health state transition diagram under Markovian assumption

SuperDrug is compared to the placebo by simulating two patient cohorts with the disease, one with the placebo treatment kernel, and the other with the estimated SuperDrug kernel 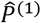. The true transition probability kernel of the placebo group is assumed well known. Meanwhile, for the SuperDrug cohort, the fictitious clinical data (*C*^(*s*)^ matrices) is generated from the true TPM 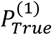. This true kernel is then blinded for the rest of the analysis.

We have described the inputs for this example in Table 1. The accrued incremental costs and quality-adjusted life years (QALYs) are used to obtain the ICER. This is repeated for each 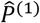 in the bootstrap. The uncertainty around the ICER is assumed to come solely from the uncertainty of 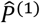 arising out of bootstrapping^11^. This was done to assess the true impact of the weighting procedure.

**Table 1:** Cost-Effectiveness Model inputs for SuperDrug versus Placebo group.

|  | Placebo Group | SuperDrug Group |
| --- | --- | --- |
| **Transition Probabilities** | Healthy → Healthy: 0.60 | Healthy → Healthy: 0.75 |
|  | Healthy → Unhealthy: 0.30 | Healthy → Unhealthy: 0.20 |
|  | Healthy → Death: 0.10 | Healthy → Death: 0.05 |
|  | Unhealthy → Healthy: 0.10 | Unhealthy → Healthy: 0.05 |
|  | Unhealthy → Unhealthy: 0.70 | Unhealthy → Unhealthy: 0.80 |
|  | Unhealthy → Death: 0.20 | Unhealthy → Death: 0.15 |
| **Total Costs (£/month)** | Healthy: 100.00<br>Unhealthy: 200.00<br>Death: 0 | Healthy: 500.00<br>Unhealthy: 600.00<br>Death: 0 |
| **Generated Input Clinical Data for algorithm** | Not applicable since true probabilities assumed to be known | $C^{(1)}: \begin{bmatrix} 186 & 71 & 13 \\ 5 & 91 & 11 \\ 0 & 0 & 138 \end{bmatrix}$ $C^{(2)}: \begin{bmatrix} 13 & 7 & 1 \\ 2 & 17 & 13 \\ 0 & 0 & 4 \end{bmatrix}$ $C^{(3)}: \begin{bmatrix} 28 & 20 & 6 \\ 5 & 20 & 18 \\ 0 & 0 & 7 \end{bmatrix}$ |
| **Health-state utilities** | Healthy: 1.00, Unhealthy: 0.7, Death: 0 |  |
| **Time Horizon** | 20 years (240 time steps) |  |

The adjustment of the weights 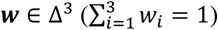 is done via a basic grid search of the 3-simplex of possible weights. The algorithm starts with uniform weights and proposes small adjustments. The variance of the ICER is recomputed for each proposed change over 5,000 bootstrap samples. A new set of weights is accepted if it reduces the variance. If none of the proposed changes are accepted, the step size is halved, and the algorithm continues. The algorithm stops when the step size reaches the required level of precision, i.e. 0.01.

Based on 5,000 samples, the proposed method resulted in a 15.2% drop in the ICER variance and a 3-percentage point increase (see Figure 3) in the probability of being cost-effective at £20,000/QALY willingness-to-pay threshold, with similar ICER means (see Table 2). Note that this method will not necessarily yield a more cost-effective estimate, it just so happens that in this example the optimization concentrates the samples around a cost-effective mean.

**Table 2:**
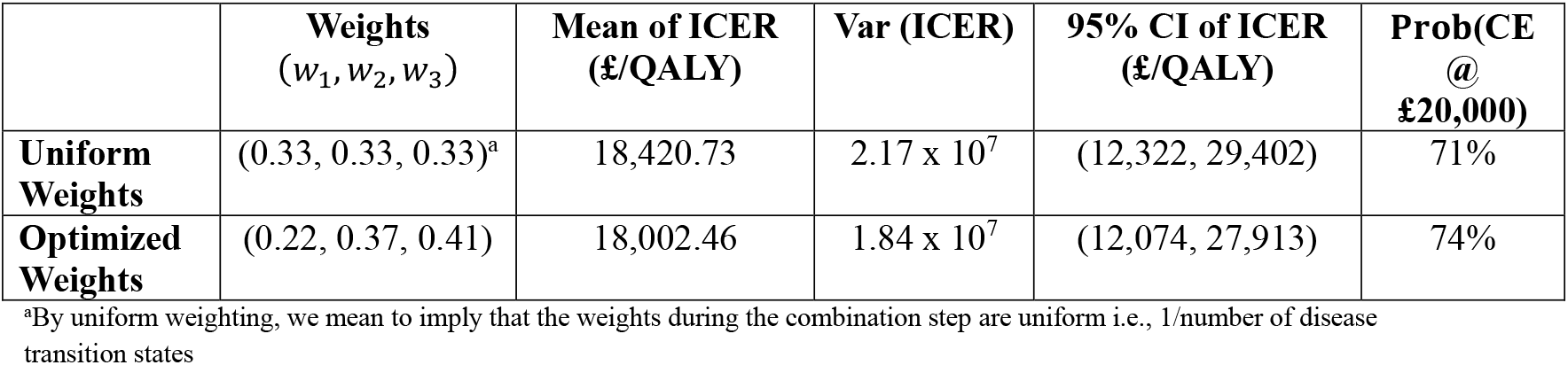
Undiscounted Cost-Effectiveness Results for SuperDrug versus Placebo.

| | **Weights**<br>($w_1, w_2, w_3$) | **Mean of ICER**<br>(£/QALY) | **Var (ICER)** | **95% CI of ICER**<br>(£/QALY) | **Prob(CE @**<br>**£20,000)** |
| --- | --- | --- | --- | --- | --- |
| **Uniform Weights** | (0.33, 0.33, 0.33)<sup>a</sup> | 18,420.73 | $2.17 \times 10^7$ | (12,322, 29,402) | 71% |
| **Optimized Weights** | (0.22, 0.37, 0.41) | 18,002.46 | $1.84 \times 10^7$ | (12,074, 27,913) | 74% |

<sup>a</sup>By uniform weighting, we mean to imply that the weights during the combination step are uniform i.e., 1/number of disease transition states

**Figure 3:**
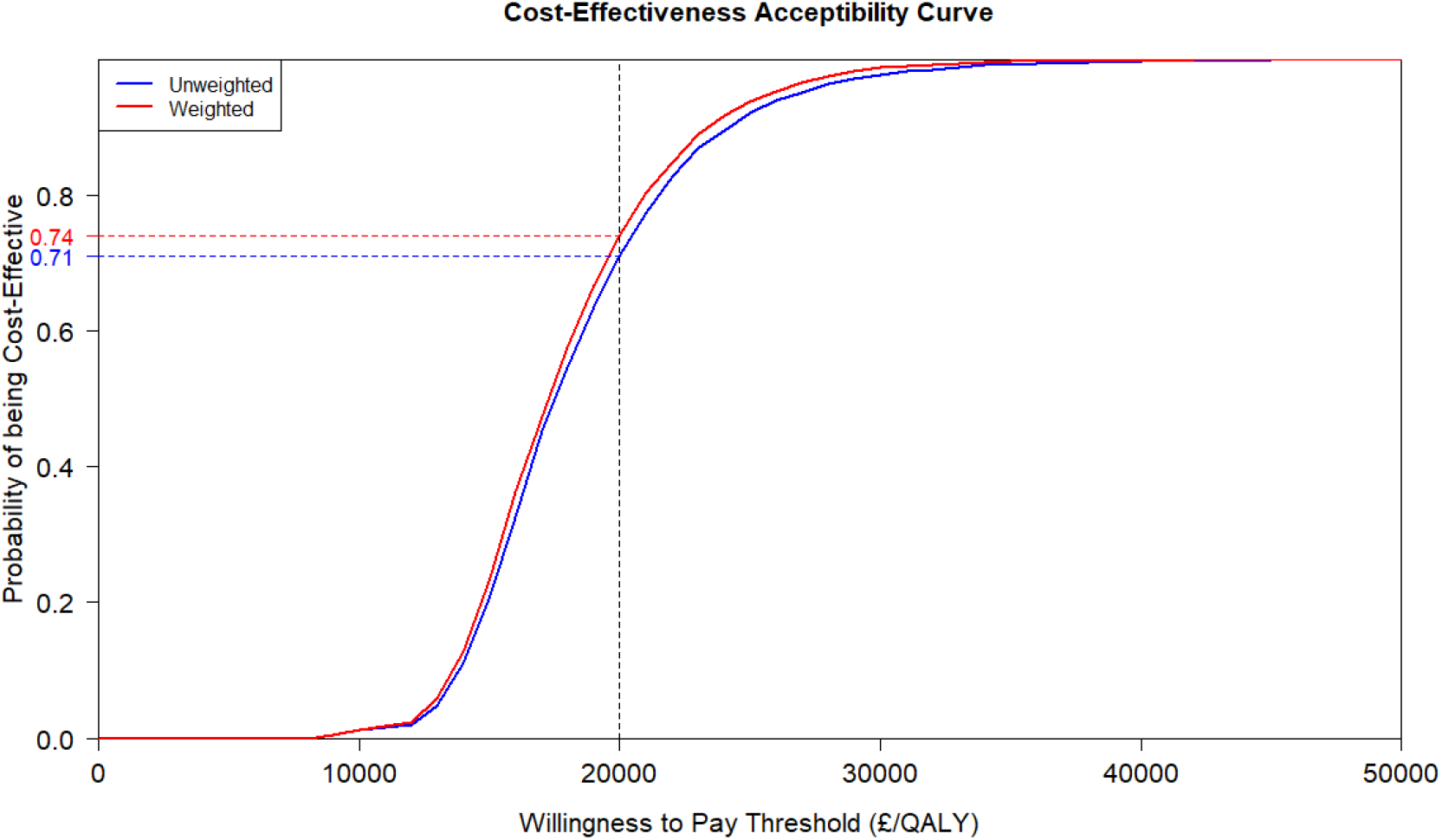
Cost-effectiveness acceptability curve for SuperDrug vs placebo

## 4. Discussion

As compared to the original uniform weighting approach, we were able to demonstrate the feasibility of using an optimized algorithm without introducing any significant estimation bias. It should be noted that our method may be viable only if each of the *C*^(*s*)^ are calculated from the same or similar studies to avoid estimation bias. However, it may happen that the analyst has to pool multiple sources of data with different sets of inclusion/exclusion criteria to arrive at the *C*^(*s*)^ matrices, resulting in a different TPM compared to using a single source. In some cases, TPMs may not be uniquely estimable. We recommend a stepwise approach when this is the case. First, the feasibility of pooling from multiple sources should be considered if one wishes to use the EM algorithm. Second, both methods should be applied and checked for any precision gain without compromising accuracy. Thirdly, the face validity of the weights should be checked with key opinion leaders if deemed necessary. Finally, one of them should be chosen as a base-case analysis with the other being used for scenario analysis. The new method results in a relatively small change in variance in our toy example, however more research is needed to determine whether a more significant change could be observed in real world clinical data. Achieving precision without introducing bias is always the target^12^, especially in HTA decision-making. For example, in rare diseases, sparse datasets with missingness are common which make it challenging for estimation. At this juncture, our method can be used to provide well-behaved PSA results along with all the benefits of using the EM algorithm in the first place. We believe that given highly variable count matrices, the fine difference between the two methods would be more prominent and visible. Naturally, the next step should be robust statistical validation by using data from large clinical trials/real-world sources.

## Data Availability

All data produced are available online at https://github.com/MarcBollee/EMforMarkov

## Acknowledgements

The authors would like to thank Subrata Bhattacharyya and Ronan Mahon for their insightful comments which helped us improve the manuscript further. We also thank Saswata Paul Choudhury for reviewing the code and Sekhar Kumar Dutta for his editorial review.

